# Post COVID-19 1^st^ Dose Vaccination Common Symptoms among SE Asia College Students

**DOI:** 10.1101/2021.12.21.21268173

**Authors:** Natasya O. Yostyadiananda, Gabriella R.A. Gunawan, Andri Wibowo

## Abstract

In the second year of the COVID-19 epidemic in the Southeast Asia (SE) regions, there is a plan to reopen the school, including the campus. Among students in Indonesia, college students have a population of almost 8.3 million. Considering the massive numbers of college students, school reopening should be supported by adequate COVID-19 vaccination. As a result, the first dose of the inactivated virus COVID-19 vaccine has been administered, including to college students aged over 18 years old. While COVID-19 vaccination is widely available, there is still a scarcity of information on post-vaccination symptoms. As reported from other locations, post vaccination has been reported. Then, this study aims to assess the common symptoms of COVID-19 1^st^ dose vaccinations among the following groups: gender (male and female college students), age, body weight, and height. The observed symptoms include sore arms, fatigue, headache, fever with a body temperature above 38 °C, nausea, shivering, and muscle joint pain. Participants in this study were students at the university. They were considered eligible for this study if they were currently enrolled at university, were at least 19 years of age, and provided informed consent. The data was recorded using a standardized online questionnaire. The answers were collected in an online database. At the beginning of the questionnaire, subjects or students were informed that data would be collected anonymously. Based on the results, the symptoms were different between female and male students. In fact, female students have experienced more symptoms than male students. While male students only suffered sore arms (68%) followed by headache symptoms (32%). Similar to male students, sore arms are the most common symptom observed among female students. Among female students, from the most to the least common symptoms observed from 20 years of age in this study are sore arms at site reaction > headache > fatigue > fever > muscle joint pain > shivering > nausea. A higher risk of presenting fatigue and headache symptoms was found in those with a non-overweight status with weight ranges of 50-60 kg.

## 1. Introduction

### 1.1 Post COVID-19 vaccination symptoms

The coronavirus SARS-CoV-2 caused the novel infectious disease COVID-19 (Liu et al 2021). Since the beginning of 2020, the disease has evolved into a global pandemic. While some patients may experience mild to moderate infection, severe cases may experience severe dyspnea, respiratory failure, and death (Dixon et al 2021). In addition to behavioral interventions such as social distancing, newly developed vaccines, which will be available in the second half of 2020, are the most important countermeasures to prevent the virus’s spread. According to available data, the vaccines approved by regulatory authorities to date have a favorable efficacy and side-effect profile. There are currently 66 vaccines in clinical trials and 176 in pre-clinical development around the world.

Aside from its efficacy, several post-vaccination symptoms have recently been observed and reported. Injection site reactions (84.1%), fatigue (62.9%), headache (55.1%), muscle pain (38.3%), chills (31.9%), joint pain (23.6%), and fever are the most commonly reported adverse events in vaccinees over the age of 16 (14.2%). These are typically mild to moderate in nature and resolve within a few days of vaccination (Pollack et al 2020).

According to recent data, the most common neurological symptom is headache, which affects more than half of those who have been vaccinated. The clinical characteristics of this headache have not yet been described in detail. In terms of headache frequency, it is known that after the first dose of vaccination, mild headaches occur in 27.4% of people, moderate headaches in 13.4% of people, and severe headaches in 1.0% of people. Mild headaches occur in 25.6% of people after the second vaccination, moderate headaches in 22.9%, and severe headaches in 3.2%.

### 1.2 College student vaccinations

In the second year of the COVID-19 pandemic, it is necessary to begin reopening educational facilities, including the campus. While young adult vaccination is an important component in reducing morbidity and mortality, there are currently 8.3 million college students. The success of vaccinations among these college students will determine the success of COVID-19 mitigation in educational facilities (Kecojevic et al. 2021).

Barello et al (2020) Pastorino et al (2020) investigated the attitudes and intentions of emerging college students toward the COVID-19 vaccine. In Italy, a high proportion of college students expressed a desire to be vaccinated. In the United States, a study of nursing faculty and students found that only 45% of students planned to get vaccinated. Another study of medical students conducted by Lucia et al (2020) discovered that 23% were unwilling to receive the COVID-19 vaccine as soon as it was approved by the Food and Drug Administration (FDA). In contrast, Graupensperger et al (2021) discovered that students regarded COVID-19 vaccination as more important than influenza vaccination in a study at a large public university in the northwest United States.

### 1.3 Inactivated virus vaccine

The most common vaccine administered is the inactivated virus vaccine. The advantages of the vaccine are that it can be stored at a normal refrigeration temperature of 2 to 8 °C and its effectiveness may last up to three years.

This vaccine is also administered in a two-dose schedule with a minimum of 14 days between each dose. The vaccine has an efficacy of 51% and is suitable for those aged 18 years and older and falls into college student criteria.

It uses inactivated vaccine technology. It means it utilizes unreactive coronavirus particles that have been killed to stimulate our bodies to produce antibodies as an immune response. This method of using an inactivated virus is a common method used to develop vaccines, which explains why this vaccine is preferred in Southeast Asia. Other vaccines that use similar methods include the polio vaccine, the Hepatitis A vaccine, and the vaccine against rabies.

### 1.4 Research significance

Despite growing studies of vaccination willingness among college students, while a few studies have also investigated post COVID-19 vaccination symptoms especially among particular college students in SE Asia. This study is very important and required immediately since potential COVID-19 cases in SE Asia an at the same time there is a plan to reopen the campus involving massive student populations

## 2. Methods

Methodology for this study is following Islam et al (2021), Mant et al (2021), and Nicolo et al (2021).

### 2.1. Participants

Participants were students at the University of Indonesia in Depok, West Java. Students were eligible if they were currently enrolled at university, were at least 19 years of age, and provided informed consent.

### 2.2. Procedure

The data related to demography, COVID-19 vaccination, and related symptoms and variables were recorded using a standardized online questionnaire. The answers were collected in an online database. At the beginning of the questionnaire, subjects or students were informed that data would be collected anonymously.

The questions are divided into the following groups: The first group was personal data on gender, age, body weight, and height (Iguacel et al 2021), followed by the type of vaccine used. In this case, the vaccine was an inactivated virus vaccine, and occurrences of post-first vaccination symptoms included sore arms, fatigue, headache, fever with a body temperature above 38 ^0^C, nausea, shivering, and muscle joint pain. Data collection was implemented from June to August 2021, when the first dose of vaccination was given and administered.

### 2.3. Data analysis

X^2^ analysis was used to assess the effects and differences of post vaccination symptoms according to gender, age, body weight, and height class. A Paired t test was used to assess the significant difference in age, height, and weight among gender groups, with a significant level at P < 0.05.

## 3. Results and Discussion

### 3.1 Demography

The demography of observed college students can be observed in Table 1. The demography was skewed because female student populations outnumbered male student populations. The number of female students was almost four times greater than that of male students. Regarding age, the male and female students were all the same age. This is supported by statistical analysis that shows fewer differences between male and female students. Those students were 20 years old on average.

**Table 1.** Demography of observed college students

| Variables | Value | P |
| --- | --- | --- |
| Sex |  |  |
| Male | 13.88% | na |
| Female | 86.11% |  |
| Age (year) |  |  |
| Male | 20.33 (95%CI: 19.7 to 21) | 0.462 |
| Female | 20.12 (95%CI: 19.6 to 20.6) |  |
| Height (cm) |  |  |
| Male | 165 (95%CI: 160 to 170) | 0.032* |
| Female | 158 (95%CI: 156 to 160) |  |
| Body weight (kg) |  |  |
| Male | 58.33 (95%CI: 53.2 to 63.4) | 0.599 |
| Female | 55.16 (95%CI: 51.1 to 59.2) |  |

\*Significant differences at $P < 0.05$

Regarding the height of students, the average height of male students was 165 cm (95%CI: 160 to 170) and female height averages were lower at 158 cm (95%CI: 156 to 160). The statistical analysis shows there was a significant difference (P < 0.05) in height averages between female and male students, where males were taller than females.

The weight averages of male students were 58.33 kg (95%CI: 53.2 to 63.4) and female weight averages were lower at 55.16 kg (95%CI: 51.1 to 59.2). The statistical analysis shows there was no significant difference (P = 0.599) in weight averages between female and male students. Actually, males and females had similar weights.

### 3.2 Post COVID-19 1^st^ dose vaccination symptoms

The most common post-COVID-19 vaccination symptoms among college students can be seen in Figure 1. There are seven types of symptoms that have been observed, including sore arms, headaches, muscle joints, shivering, nausea, fatigue, and fever. Obvious symptom differences observed were that female students experienced more symptoms than male students. While male students only suffered sore arms and were followed by headache symptoms. Similar to male students, sore arms are the most common symptom observed among female students.

**Fig 1.**
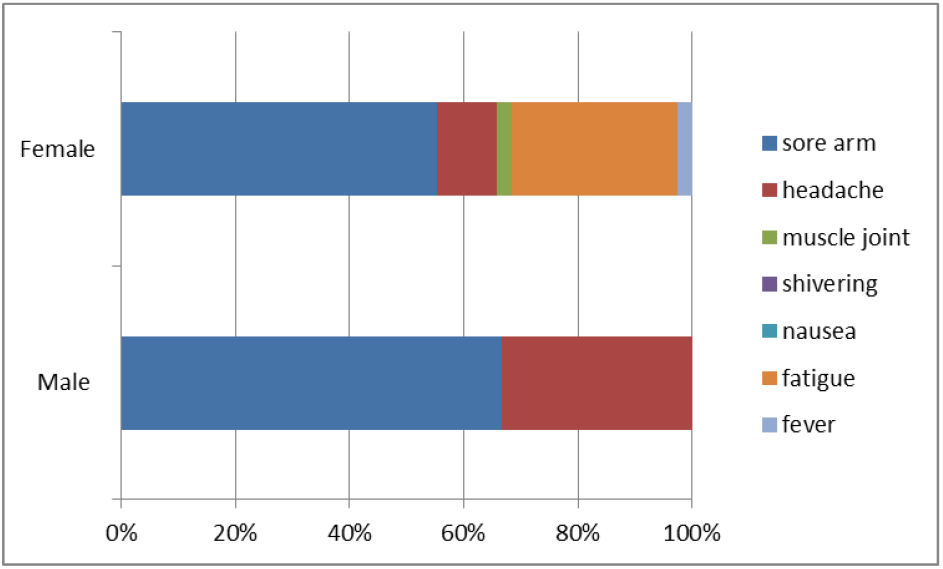
Compositions of common post COVID-19 1^st^ dose vaccination symptoms between male and female college students

Figure 2 shows in more detail the frequencies of each symptom among female students according to age, weight, and height. Female students aged 20 years old mostly experience sore arms, followed by fatigue and headaches. Sore arms were most commonly observed among female students with a height class range of 150–162 cm. Other symptoms observed in this height class were fatigue, followed by headache. In contrast, female students with a height class range of 165–170 cm had fewer symptoms, and even headache syndrome was absent in this group.

**Fig 2.**
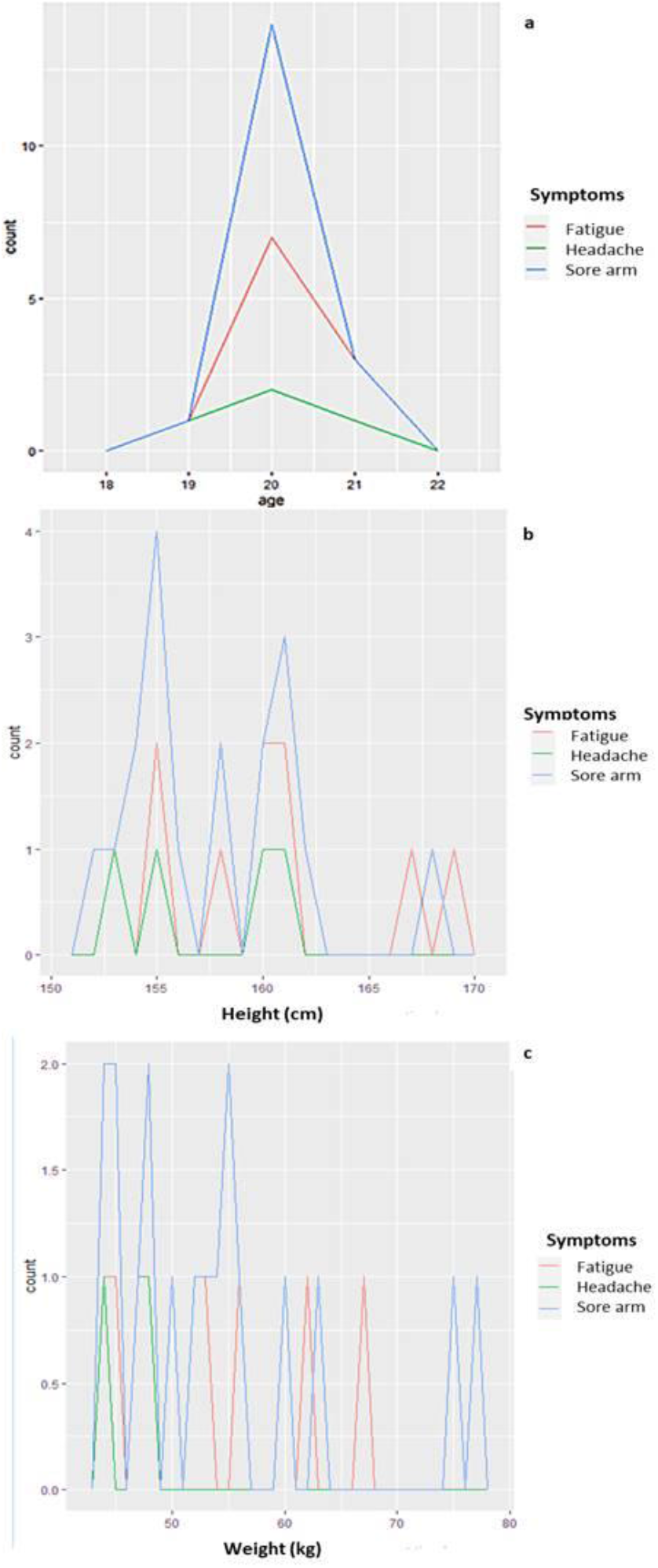
Post COVID-19 1^st^ dose vaccination symptom distributions based on age, height (cm), and weight (kg) classes among female college students

Similar patterns of symptoms were also observed in the weight class. Female students with a weight class range of 50–60 kg have experienced more sore arm symptoms. Within this class, fatigue and headache symptoms occurred at the same frequencies. Tall female students with heights greater than 170 cm, on the other hand, only experience sore arm and fatigue symptoms. In this class, headache was absent.

### 3.3. Discussion

The post-COVID-19 symptoms observed in this study were in agreement with other results. According to Gobel et al. (2021), the most frequently reported adverse events in vaccinees over the age of 16 are injection site reactions (84.1%), fatigue (62.9%), headache (55.1%), muscle pain (38.3%), chills (31.9%), joint pain (23.6%), and fever (14.2%). In this study, the most common symptoms observed from 20 years of age were sore arms at site reaction > headache > fatigue > fever > muscle joints, shivering > nausea. Then it seems that sore arms, fatigue, and headaches are the most common post-COVID-19 symptoms. According to Vaccaro et al 2021, more than 80% of vaccinees have developed injection site pain, followed by erythema and swelling cases that were present in about 10%. This is also consistent with findings by Chapin-Bardales et al (2021) that found that injection site pain is common after both the first and second doses of either vaccine. Those symptoms are considered systemic reactions, including fatigue, headache, myalgia, chills, fever, and joint pain, and usually occur in participants after the first dose.

This study observed significant occurrences of headache symptoms. The presence of this symptom was different between male and female students. Consistent with this finding, Gobel et al. (2021) have also reported a significant difference between women and men with regard to the intensity of headache attributed to COVID-19 vaccination (3.4 ± 0.8 versus 3.1 ± 0.8; P < 0.001). Headaches after the COVID-19 vaccination show an extensive complex of symptoms. Headaches occur with similar symptoms in all age and gender groups. Less than 10% of those affected also report headaches resulting from previous vaccinations. The headache after a COVID-19 vaccination occurs on average around 18 h after vaccination. It lasts an average of 14 hours. In around two-thirds of those affected, the headache lasts between 1 and 12 hours. Over 66% experienced a monophasic headache course. The headache usually occurs bilaterally, with the main locations being the forehead, temples, back of the head, and retro orbital region. They do not radiate to other regions. The pain character is most frequently described as being pressurized and dull. The pain intensity is moderate to severe.

Routine physical activity can exacerbate the headache. This is consistent with our findings, as male college students were more active than female students. In fact, male students were doing more sports include soccer and running. The most common accompanying symptoms are fatigue, exhaustion, and muscle pain. These can be accompanied by nausea as well as hypersensitivity to light and noise. In women, the duration and intensity of headaches are significantly greater than in men. This analysis does not aim to attribute a cause to each individual symptom. The aim of the study is to record the headache symptom complex in temporal relation to the vaccination. With regard to the vaccine type used for the very first time, the aim was to determine this complex of symptoms in detail.

The presences of headache symptoms may be related to the type of vaccine used in this study that is inactivated virus vaccine. The association of inactivated virus vaccine has been highlighted by Ekizoglu et al 2021. An inactivated viral vaccine is containing alum, an aluminum salt–based adjuvant. A current study by Pulendran and Ahme (2011) have demonstrated that alum signals through the NLRP3 inflammasome leading to caspase 1-dependent release of the pro-inflammatory cytokines IL-1β and IL-18.

## 4. Conclusions

At the time of this study and to the best of our knowledge, no research had examined post-COVID-19 1^st^ dose inactivated virus vaccination symptoms among more diverse college student populations. To conclude, there is a discrepancy in symptoms between males and females. The weight and height of the vaccinees, especially female college students, were also contributing to the symptoms.

## Data Availability

All data produced in the present work are contained in the manuscript

